# Progressive changes in glutamate concentration in early stages of schizophrenia: A longitudinal 7-Tesla MRS study

**DOI:** 10.1101/2020.08.11.20172841

**Authors:** Peter Jeon, Roberto Limongi, Sabrina Ford, Michael Mackinley, Kara Dempster, Jean Théberge, Lena Palaniyappan

## Abstract

Progressive reduction in glutamatergic transmission has been proposed as an important component of the illness trajectory of schizophrenia. Despite its popularity, to date, this notion has not been convincingly tested in patients in early stages schizophrenia. In a longitudinal 7T magnetic resonance spectroscopy (1H-MRS), we quantified glutamate at the dorsal anterior cingulate cortex in 21 participants with a median lifetime antipsychotic exposure of less than 3 days and followed them up after 6 months of treatment. Healthy controls were also scanned at two time points. While patients had significantly lower overall glutamate levels than healthy controls (F(1,27) = 5.23, p = 0.03), we did not observe a progressive change of glutamate concentration in patients (F(1,18) = 0.47, p = 0.50), and the group by time interaction was not significant (F(1,27) = 0.86, p = 0.36). On average, patients with early psychosis receiving treatment showed a 0.02 mM/year increase, while healthy controls showed a 0.06 mM/year reduction of MRS glutamate levels. Bayesian analysis of our observations does not support early, post-onset glutamate loss in schizophrenia. Interestingly, it provides evidence in favour of a lack of progressive glutamate change in our schizophrenia sample – indicating that the glutamatergic level at the onset of illness was the best predictor of the levels 6 months after treatment. A more nuanced view of glutamatergic physiology, linked to early cortical maturation, may be required to understand glutamatergic dynamics in schizophrenia.

## Introduction

Glutamatergic disruption is implicated in the wide range of symptoms observed in schizophrenia ^1–3^. Specifically, disinhibition of the excitatory glutamatergic outputs of the prefrontal cortex is thought to disrupt dopaminergic signaling in the striatum ^4,5^ resulting in acute psychotic symptoms. However, sustained disinhibition of prefrontal glutamatergic neurons might lead to excitotoxic damage with subsequent reduction in glutamate, with greater reductions occurring in patients with more severe forms of schizophrenia ^6^. Magnetic resonance spectroscopy (MRS) studies in schizophrenia report higher glutamate levels in younger patients at early stages while lower glutamate levels in older patients at later stages of illness, when compared to healthy controls ^7^.

A progressive pathology defined by gray matter changes ^8–10^, ventricular enlargement ^11–15^, and network-level dysconnectivity ^16,17^ is thought to be the basis of the longitudinal trajectory of schizophrenia. It is posited that glutamatergic dendritic spine reduction triggered by early excitotoxic processes lies at the centre of such morphological changes ^18,19^. The progressive pathology of schizophrenia is likely limited to certain hubs of the brain ^16^, with the anterior cingulate cortex (ACC) being a prominent region where both structural, functional ^20^ and neurochemical deficits ^21–24^ have been consistently demonstrated in schizophrenia. Nevertheless, longitudinal MRS studies investigating progressive glutamate changes in the ACC are limited.

Using 1.5T in patients at various stages of schizophrenia, Choe et al. ^25^ reported a notable reduction in prefrontal Glx (glutamate + glutamine) signal 1-6 months after treatment. Théberge et al. ^10^ and Bustillo et al. ^26^ demonstrated static glutamate levels in the ACC using 4T. Merritt et al. ^27^ also failed to see a progressive reduction in Glx using 3T in schizophrenia. When using 7T MRS, with superior specificity for glutamate quantification ^28^, a cross-sectional association of decreasing ACC glutamate with increasing age was observed in schizophrenia ^29^ at an accelerated rate compared to healthy young adults ^30^. Nevertheless, to date, longitudinal 7T MRS studies have not been reported in schizophrenia.

In this study, we tested if (1) glutamatergic deficit indexed by ACC MRS measure of glutamate is present in early stages of psychosis, (2) whether this deficit progressively worsens in the first 6 months of treatment, and (3) if patients show an exaggerated longitudinal decline compared to healthy controls. To our knowledge, this is the first longitudinal report of 7T MRS in schizophrenia.

## Methods

### Participants

We recruited 21 first-episode schizophrenia (FES) volunteers with inclusion criteria of lifetime antipsychotic exposure being less than 14 days along with 10 healthy control volunteers, group-matched for age, gender, and parental socio-economic status. Patient volunteers were recruited from the referrals received by the PEPP (Prevention and Early Intervention for Psychosis Program) at London Health Sciences Center. All patients had established consensus diagnosis after 6 months of first-episode schizophrenia by 3 psychiatrists (LP, KD, and primary treatment provider at PEPP) based on the DSM-5 criteria ^31^. Participants whose 6-month diagnoses were bipolar or major depressive disorder with psychoses as well as suspected drug-induced psychoses were excluded from the study. Healthy control volunteers had no personal history of mental illness and no family history of psychotic disorder. All participants were screened to exclude significant head injury, major medical illness, or MRI contraindications and provided written, informed consent according to the guidelines of the Human Research Ethics Board for Health Sciences at Western University, London, Ontario.

### MRS Acquisition and Analysis

MRS measurements were acquired using a Siemens MAGNETOM 7T head-only MRI scanner (Siemens, Erlangen, Germany) and a site-built head coil (8-channel transmit, 32-channel receive) at the Centre for Functional and Metabolic Mapping of Western University (London, Ontario). A two-dimensional sagittal anatomical image (37 slices, TR = 8000 ms, TE = 70 ms, flip-angle (*α*) = 120°, thickness = 3.5 mm, field of view = 240×191 mm) was used as reference to prescribe a 2.0 × 2.0 × 2.0 cm (8 cm^3^) ^1^H-MRS voxel on the bilateral dorsal ACC (Figure 1). Voxel positioning was set by having the posterior end of the voxel coinciding with the precentral gyrus and the caudal face of the voxel coinciding with the most caudal positioning that was not part of the corpus callosum. Voxel angle was set to be tangential to the corpus callosum. A semi-LASER ^1^H-MRS sequence (TR = 7500 ms, TE = 100 ms, bandwidth = 6000 Hz, N = 2048) was used to acquire 32 channel-combined, VAPOR ^32^ water-suppressed spectra as well as a water-unsuppressed spectrum to be used for spectral editing and quantification. During scan, participants were asked to rest by fixing their gaze on a white cross on a 50% gray background.

**Figure 1.**
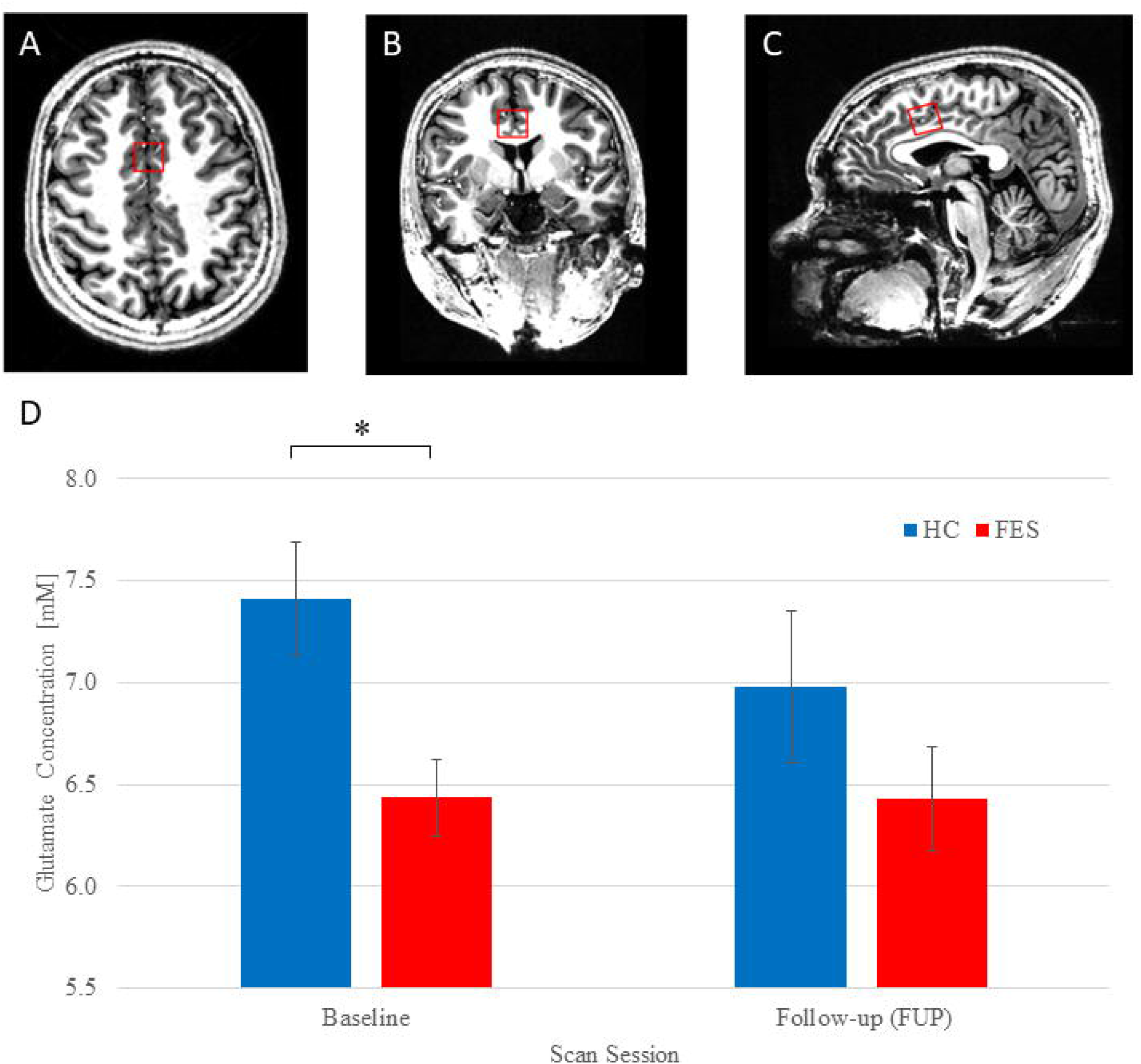
(A) Axial, (B) coronal, and (C) sagittal views of MRS voxel (red square) in the dorsolateral anterior cingulate cortex (ACC) for glutamate measurement. (D) Estimated marginal means of glutamate concentration [mM] for healthy controls (HC, blue) and patients (FES, red) at baseline and follow-up scan sessions. Error bars indicate standard deviation. Asterisk denotes significant difference. (*Note: y-axis values do not begin at 0 for graphical purposes*).

Using the tools outlined in Near et al. ^33^, the 32 spectra were phase and frequency corrected before being averaged into a single spectrum to be used for all subsequent analyses. QUECC ^34^ and HSVD ^35^ were applied to the spectrum for lineshape deconvolution and removal of residual water signal, respectively. Spectral fitting was done using fitMAN ^36^, a time-domain fitting algorithm that uses a non-linear, iterative Levenberg-Marquardt minimization algorithm to echo time-specific prior knowledge templates. The metabolite fitting template included 17 brain metabolites: alanine, aspartate, choline, creatine, γ -aminobutyric acid (GABA), glucose, glutamate, glutamine, glutathione, glycine, lactate, myo-inositol, N-acetyl aspartate, N-acetyl aspartyl glutamate, phosphorylethanolamine, scyllo-inositol, and taurine. No significant macromolecule contribution was expected due to the long echo time and hence was omitted from the metabolite template. Metabolite quantification was then performed using Barstool ^37^ with corrections made for tissue-specific (gray matter, white matter, CSF) T_1_ and T_2_ relaxations through partial volume segmentation calculations of voxels mapped onto T_1_-weighted images acquired using a 0.75 mm isotropic MP2RAGE sequence (TR = 6000 ms, TI_1_ = 800 ms, TI_2_ = 2700 ms, flip-angle 1 (*α*_1_) = 4°, flip-angle 2 (*α*_2_) = 5°, FOV = 350 mm × 263 mm × 350 mm, T_acq_ = 9 min 38 s, iPAT_PE_ = 3 and 6/8 partial k-space). All spectral fit underwent visual quality inspection as well as Cramer-Rao lower bounds (CRLB) assessment for each metabolite.

### Clinical Assessments

Symptom severity at baseline was measured using PANSS-8 ^38^ scale, on the same day of the first scan. We also quantified the overall social and occupational functioning at the time of first presentation using SOFAS ^39^, administered on the same day of the scanning. We assessed Duration of Untreated Psychosis (DUP) based on multiple sources of information provided by the patient, the referring sources, and caregivers as well as by reviewing clinical charts. We used the first emergence of positive psychotic symptoms as the starting point for calculating the DUP, in line with prior work in this regard ^40^.

To determine cannabis use in the past six months, the Cannabis Abuse Screening Test (CAST) was used ^41^. The CAST is a six-item Likert-scale self-report questionnaire which asks the participant about cannabis use and how it effects their daily activities and relationships. Scores range from six to 30, with higher scores indicating more cannabis use. To determine alcohol use in the past six months, the Alcohol Use Disorders Identification Test (AUDIT-C) ^42^ was used. The AUDIT-C is a three-item Likert-scale self-report questionnaire which asks the participant about alcohol use frequency and quantity. Scores range from 0 to 12, with higher scores indicating more alcohol use. Alcohol users and nonusers were classified by AUDIT-C scores of four or more and less than four, respectively. Lastly, nicotine use in the past six months was determined by the single item Fagerström Test for Nicotine Dependence and smoking index ^43^. The Fagerström test indicates time to the first cigarette after waking, and the smoking index is calculated by the number of years regularly smoking × the number of cigarettes per day, divided by 20, to determine packs per year. A lower Fagerström test value indicates more nicotine dependence, and a higher smoking index indicates more nicotine use. The 10-item Drug Abuse Screening Test (DAST-10) ^44^ was also employed for substances other than cannabis, alcohol and nicotine, though our cohort did not endorse any such use.

### Statistical Analyses

All frequentist statistical tests were computed using IBM SPSS Statistics version 26 ^45^. Group demographic differences were calculated using *t* tests and chi-square tests for continuous and dichotomous variables, respectively. Repeated measure ANOVA was used to assess group × time interaction (primary hypothesis), as well as group effect and time effect, with parameter estimates examined to test individual group effects. As age and gender are known modifiers of glutamate levels, they were entered as covariates in the ANOVA model. Lastly, Pearson correlation was used to explore the correlation of annualized, baseline-adjusted glutamate change to DUP, SOFAS, as well as symptom severity at first presentation, measured using PANSS-8 total score at baseline in patients. Correlations between defined daily dose (DDD) and annualized, baseline-adjusted glutamate change and follow-up glutamate concentrations were also examined.

To investigate relationships of annualized glutamate with cannabis and nicotine, Pearson correlations were used. Glutamate change was analyzed with total CAST to determine relationships with cannabis, along with smoking index and Fagerström scores to determine nicotine use. To determine alcohol use, a *t*-test was used to compare glutamate change values between alcohol users and nonusers.

We performed a (Bayesian) hierarchical generalization of an analysis of covariance (Bayesian ANCOVA) to evaluate whether there is a between-group difference of the effect of time on the follow-up measurement of glutamate concentration. We decided to use a Bayesian approach as an alternative to traditional frequentist test because it allows us to weigh the evidence in support of our main hypothesis relative to the evidence in support of the null hypothesis. We achieved this via model comparison and Bayes factors (BF) ^46,47^.

We fit one saturated ANCOVA model and all possible reduced models comprising follow-up glutamate concentration ([Glu]_follow-up_) as a dependent variable, Group as factor, and baseline glutamate concentration ([Glu]_baseline_) and Interval (days) as covariates. We compared the evidence supporting this (saturated) model with the evidence supporting the reduced models (including the null model). We relied on the largest BF10 relative to the null model to select the winning model. Note that we included [Glu]_baseline_ as a covariate of no interest. Therefore, our main hypothesis was represented by the triple-interaction model (Group × [Glu]_baseline_ × Interval).

In all models, the posterior distributions over parameters were estimated using the ‘generaltestBF’ function in the “R Bayes Factor” package ^48^. In this data set, the small number of subjects in each group might cause the posterior distribution to be strongly influenced by the prior distribution. Therefore, we used informed “wide” priors scaled to the observed data (r-scale for each effect = 0.5). We report the mean and standard deviation for each estimate obtained from the relevant posterior distribution (10,000 samples) along with the 95% highest density interval (HDI).

## Results

### Demographic Data

Demographic and clinical data of subjects are shown in Table 1. Our patient sample had a mean DUP of 29.38 weeks (SD = 26.65 weeks) and a mean antipsychotic duration of 2.95 days (SD = 3.11 days) prior to the first scan session. Patient and healthy control SOFAS scores were significantly different (t(29) = 12.466, *p* < 0.001). The time in between baseline and follow-up (FUP) scan was 5.93 months (SD = 1.25) for patients and 7.25 months (SD = 1.90) for healthy controls.

**Table 1.** Demographic and clinical characteristics.

| Characteristic | Patient group<br>(*N* = 21) | Healthy controls<br>(*N* = 10) | *t*/$\chi^2$ | *p* |
| --- | --- | --- | --- | --- |
| Gender (male/female) | 16/5 | 5/5 | 2.13 | 0.145 |
| Marital status (Mar/S) | 1/20 | 1/9 | 0.31 | 0.58 |
| Inpatient at baseline (Y/N) | 11/10 |  |  |  |
| Family Hx (Y/N/DN) | 10/6/5 |  |  |  |
| AP exposure at baseline (M/SD; days) | 2.95/3.11 |  |  |  |
| Total DDD-days at baseline scan (M/SD) | 2.25/4.74 |  |  |  |
| Total DDD-days at FUP scan (M/SD) | 145.68/97.56 |  |  |  |
| DUP (M/SD/median; weeks) | 29.38/26.65/18 |  |  |  |
| Ethnicity (Black/White/Other) | 2/18/1 | 0/5/5 | 4.51 | 0.034<sup>a</sup> |
| Age (M/SD) | 22.33/5.29 | 21.60/3.37 | -0.47 | 0.645 |
| SOFAS (M/SD) | 42.33/12.84 | 83.70/5.62 | 12.47 | 0.000 |
| PANSS-8 total (M/SD) | 24.67/5.30 |  |  |  |
| Smoker (yes/no) | 6/15 | 0/10 | 3.54 | 0.060 |
| Cannabis user (yes/no) | 13/8 | 0/10 | 10.66 | 0.001 |
| Time between scans (M/SD; months) | 5.93/1.25 | 7.67/1.90 | 2.63 | 0.021 |

*P* values for differences between groups were calculated using chi-square analyses for categorical variables and independent *t* tests for continuous variables.
*Mar* married, *S* single, *Y* yes, *N* no, *Hx* history, *DN* don't know, *AP* antipsychotic, *M* mean, *SD* standard deviation, *DDD* defined daily dose, *FUP* follow-up, *DUP* duration untreated psychosis.
<sup>a</sup> White vs non-White comparison.

CRLB values indicating the quality of glutamate measurement was quantified for both groups. For HC glutamate CRLB were 3.41% (SD = 1.27%) and 3.55 % (SD = 0.89%) for baseline and FUP, respectively. For FES glutamate quantification, CRLB values were 3.52% (SD = 1.20%) and 3.96% (SD = 1.12%) for baseline and FUP, respectively. Thus the 2 groups had acceptable qualitative metrics for glutamate estimation at both time points.

### Longitudinal Glutamate

Paired *t* tests revealed no significant differences between FES (M = 6.51 mM, SD = 0.64 mM) and HC (M = 7.25 mM, SD = 1.34 mM) unadjusted baseline glutamate concentration (t(29) = 1.66, *p* = 0.13, Cohen’s d = 0.70) as well as unadjusted FUP glutamate concentration (M = 6.49 mM, SD = 1.29 mM; M = 6.86 mM, SD = 0.73 mM; t(29) = 1.02, *p* = 0.32, Cohen’s d = 0.35). Repeated measures ANOVA revealed a group effect (F(1,27) = 5.23, *p* = 0.03, Cohen’s d = 0.90) between FES (M = 6.43 mM, SD = 0.84 mM) and HC (M = 7.20 mM, SD = 0.86 mM) but no effect on time (F(1,27) = 1.21, *p* = 0.28) or group × time interaction (F(1,27) = 0.86, *p* = 0.36). Parameter estimates revealed that at baseline, FES had lower glutamate than healthy controls (t(29) = 2.83, *p* = 0.009, Cohen’s d = 1.11), but this difference was not present at follow-up (t(29) = 1.20, *p* = 0.24, Cohen’s d = 0.41). A simple contrast of time in each group revealed no significant effect in both the healthy control group (F(1,7) = 0.25, *p* = 0.63, Cohen’s d = 0.41) and in patients (F(1,18) = 0.47, *p* = 0.50, Cohen’s d = 0.003) (Figure 1).

Annualized glutamate concentration values were not significantly different between the two groups (t(29) = -0.813, *p* = 0.423) and indicated a 0.02 mM/year (SD = 0.33 mM) increase in patients and a 0.06 mM/year (SD = 0.19) reduction in healthy controls, with the difference amounting to a small to moderate sized effect (Cohen’s d = 0.26). Lastly, the time interval between scans in months was not related to glutamate concentration differences (FUP – baseline) in either group (HC: *r* = -0.23, *p* = 0.52; FES: *r* = -0.41, *p* = 0.06).

### Glutamate versus Clinical Measures

There was no significant correlation between annualized glutamate concentration changes and DUP (*r* = -0.07, *p* = 0.77), SOFAS (*r* = -0.09, *p* = 0.70), or PANSS-8 total (*r* = 0.25, *p* = 0.28). We also did not see any correlation between baseline (unadjusted) glutamate concentration and DUP (*r* = 0.03, *p* = 0.91), SOFAS (*r* = -0.38, *p* = 0.09), or PANSS-8 total (*r* = 0.31, *p* = 0.17).

Across all participants, there was no significant difference in annualized glutamate change between alcohol users (*n* = 23) and nonusers (*n* = 6) (*t*(27) = 1.89, *p* = 0.07). No significant difference was found between alcohol users and nonusers when FES was considered (*t*(17) = 1.37, *p* = 0.19) separately. No significant difference was found in annualized glutamate change between smokers and non-smokers among the FES patients (*t*(20) = -0.72, *p* = 0.63). Across all participants, there was no significant correlation between total CAST scores (*n* = 28) and annualized glutamate change (*r* = -0.08, *p* = 0.75). Lastly, there was no significant correlation between DDD at follow-up (*n* = 21) and follow-up glutamate concentration (*r* = -0.16, *p* = 0.50) as well as between DDD at follow-up and annualized glutamate change (*r* = -0.16, *p* = 0.48).

### Bayesian Statistical Analysis

After controlling for baseline values, the [Glu]_follow-up_ in the FEP group was the same as in the HC group. The Bayesian ANCOVA model revealed that the triple interaction (Group × [Glu]_baseline_ × Interval) did not perform better than the null model (Table 2). This means that, in practice, the longitudinal change in glutamate concentration of both groups are alike.

**Table 2.** Bayesian model comparison.

| Model | $BF_{10}$ |
| --- | --- |
| **N** | 1.000 |
| **G** | 0.465 |
| **B** | **4.080** |
| $G + B$ | 1.547 |
| $G + B + G \times B$ | 1.732 |
| **I** | 0.351 |
| $G + I$ | 0.201 |
| **B + I** | **3.126** |
| $G + B + I$ | 1.451 |
| $G + B + G \times B + I$ | 1.828 |
| $G + I + G \times I$ | 0.155 |
| $G + B + I + G \times I$ | 0.905 |
| $G + B + G \times B + I + G \times I$ | 1.929 |
| **B + I + B × I** | **3.164** |
| $G + B + I + B \times I$ | 1.448 |
| $G + B + G \times B + I + B \times I$ | 0.980 |
| $G + B + I + G \times I + B \times I$ | 2.855 |
| $G + B + G \times B + I + G \times I + B \times I$ | 1.814 |
| $G + B + G \times B + I + G \times I + B \times I + G \times B \times I$ | 0.960 |

$BF_{10}$ is computed relative to null model.
$N$ null, $B$ baseline, $I$ interval, $G$ group.

Table 2 also shows that a model including only the [Glu]_baseline_, a model including both [Glu]_baseline_ and Interval, and a model including its interaction outperformed the null model (BF10 > 3). However, the more complex models underperformed the “[Glu]_baseline_” model (the best model). This suggests that, with moderate evidence (10 > BF10 > 3) ^47^, [Glu]_baseline_ is the best predictor of [Glu]_follow-up_ regardless of both follow-up measurement time and Group. Table 3 shows the relevant parameter estimates along with the 95% interval of most credible values.

**Table 3.** Model Averaged Posterior Summary

| Parameter | Mean | SD | 95% HDI |  |
| --- | --- | --- | --- | --- |
|  |  |  | Lower | Upper |
| **Intercept** | **6.617** | **0.205** | **6.209** | **7.020** |
| FEP | -0.074 | 0.178 | -0.434 | 0.270 |
| HC | 0.074 | 0.178 | -0.270 | 0.434 |
| **Baseline** | **0.511** | **0.247** | **0.048** | **1.007** |
| Interval | -0.004 | 0.004 | -0.012 | 0.003 |
| FEP × [Glu]<sub>baseline</sub> | 0.149 | 0.225 | -0.288 | 0.595 |
| HC × [Glu]<sub>baseline</sub> | -0.149 | 0.225 | -0.595 | 0.288 |
| FEP × Interval | -0.006 | 0.004 | -0.014 | 0.001 |
| HC × Interval | 0.006 | 0.004 | -0.001 | 0.014 |
| [Glu]<sub>baseline</sub> × Interval | -0.005 | 0.006 | -0.016 | 0.006 |
| FEP × Interval × [Glu]<sub>baseline</sub> | -0.002 | 0.005 | -0.013 | 0.009 |
| HC × Interval × [Glu]<sub>baseline</sub> | 0.002 | 0.005 | -0.009 | 0.013 |
| **$\sigma^2$** | **0.964** | **0.276** | **0.560** | **1.626** |

Only the Intercept, [Glu]<sub>baseline</sub>, and sig2 parameters (bolded) were granted with PP > 0.95.
$\sigma^2$ posterior variance of the “deflection parameters”, PP posterior probability.

## Discussion

We report longitudinal 7T-MRS glutamate measurements in FES patients who were medication-naïve at baseline. Although baseline glutamate concentrations were lower in FES compared to HC, follow-up measurements revealed no difference in glutamate concentration between the two groups. Our current work supports the lower glutamate concentration observed in FES compared to HC at baseline as reported in current literature ^49^. Annualized glutamate concentration changes also showed no difference between FES and HC. Bayesian statistical approach also provided evidence in favour of a lack of progressive glutamate change in our FES sample, indicating that the glutamatergic level at the onset of illness was the best predictor of the levels 6 months after treatment. Taken together, our results indicate that early in the illness, patients with schizophrenia already show abnormalities in ACC glutamate, and do not show evidence for a progressive deterioration in glutamatergic status.

Prior longitudinal studies at lower field strengths have been equivocal on the issue of progressive glutamatergic reduction in schizophrenia ^7^. Synthesizing this longitudinal literature, Egerton et al. ^50^ noted an almost even split between studies that reported significant glutamate decrease in at least one brain region and studies reported no glutamate reduction, with at least 3 further studies showing no longitudinal glutamate reduction in the ACC after 6 weeks ^27,51,52^, 4 months ^51^, and 9 months ^27^. Our findings are more aligned with these recent studies as well as the lack of progressive ACC glutamate change reported by Bustillo et al. ^26^ [at 1, 6, 12 months] and Théberge et al. ^10^ [10 months] at 4T. In contrast, Egerton et al. ^53^ observed a reduction in glutamate to creatine ratios at 3T in the ACC after 4 weeks of antipsychotic treatment, though 2 of the 3 sites in this multi-site study did not show the same effect. In summary, our current study supports the extant literature on the lack of progressive glutamate changes in the ACC during the early course of treatment in schizophrenia.

Our study has a number of strengths including the use of 7T scanner, and recruiting highly symptomatic, mostly drug-naïve individuals. Several limitations should also be considered when interpreting the results. We chose a single voxel (dorsal ACC) for the current 6-month follow-up study. We cannot exclude the possibility of progressive glutamate changes in different brain regions (as shown by Théberge et al. ^10^ and Goto et al. ^54^ in thalamus and basal ganglia) or in ACC over a longer time scale with multiple time points. Additionally, though glutamine is sometimes considered to reflect the synaptic pool of glutamate due to its glial localisation, our acquisition was not optimised for quantifying glutamine; we had high CRLB values (23.70 ± 15.93% and 26.83 ± 12.06% for baseline and follow-up, respectively) for glutamine quantification. As a result, the glutamate concentration reported here must be interpreted as reflective of largely neuronal origin ^55,56^.

Although we had sufficient power to detect if paired differences were present in the patient group, any interaction is likely underpowered. We observed a small glutamate decrease in healthy control (0.06 mM/year), comparable to the effect size observed in Birur et al. ^51^ with 16 patients and 14 healthy controls. Bayesian statistical approaches are based on an expected prior distribution and not influenced by the central limit of sample size placed on conventional statistics. Bayesian approach also confirmed the evidence in favour of a lack of progressive glutamatergic changes in schizophrenia. For obvious ethical reasons, we lacked a patient group that remained untreated for 6 months to parse the effects of treatment from illness stage.

In summary, this longitudinal 7T MRS study of ACC found a stable glutamatergic deficit that does not progressively worsen in the early stages of schizophrenia. This supports the possibility of the putative excitotoxic processes predating the first presentation of psychosis, either in the prodromal stages or more distally during early development. Further, this also challenges the notion of a relentlessly progressive glutamatergic dysfunction in patients receiving treatment.

## Data Availability

Data is available upon request. Please email the corresponding author.

## Acknowledgements

We thank Mr. Trevor Szekeres, Mr. Scott Charlton, Mr. Joseph Gati for their assistance in data acquisition and archiving. We thank all research team members of the NIMI lab and all the staff members of the PEPP London team for their assistance in patient recruitment and supporting clinical care. We gratefully acknowledge the participants and their family members for their contributions. Requests for data should be addressed to Dr. Lena Palaniyappan.

